# Re-evaluation Of Hypo- And Hyperoxemia In Patients With Respiratory Failure And Veno-Venous Extracorporeal Membrane Oxygenation

**DOI:** 10.64898/2026.04.01.26349732

**Authors:** Victoria Bünger, Martin Ruß, Luigi La Via, Oliver Hunsicker, Mario Menk, Wolfgang M. Kuebler, Steffen Weber-Carstens, Jan A. Graw

## Abstract

**Background:** Many patients in the ICU receive oxygen to secure blood and tissue oxygenation. Increasing evidence shows exposure to high fractions of inhaled oxygen (FiO_2_) being associated with adverse effects. In patients with severe ARDS, veno-venous Extracorporeal Membrane Oxygenation (VV-ECMO) can be implemented as a rescue therapy and PaO_2_ levels can be controlled by the blood flow of the VV-ECMO. Yet, optimal oxygenation targets in ARDS patients treated with VV-ECMO are unknown.

**Methods:** Retrospective analysis of 443 patients with severe ARDS treated with VV-ECMO. Regression analyses were performed for mortality and time-weighted averages of PaO_2_ and FiO_2_. Furthermore, considering a possible non-linear relationship, a restricted cubic spline (RCS) model was performed for PaO_2_.

**Results:** A simple logistic regression for mean PaO_2_ and ICU mortality showed a significant positive association (per mmHg OR 0.99 [95%CI 0.98-1.00], p=0.002). RCS analysis showed a U-shaped association of mortality and mean paO_2_ (paO_2_ 69.70-90.24mmHg: OR 0.92 [95%CI 0.89-0.94], p<0.001; paO_2_ 90.24-123.40mmHg: OR 1.09 [95%CI 1.06-1.13], p<0.001). A model including PaO_2_ as RCS variable and FiO_2_ showed significant associations of mortality with both variables (PaO_2_ 69.70-90.24mmHg: OR 0.94 [95%CI 0.91-0.97], p<0.001; paO_2_ 90.24-123.40 mmHg: OR 1.07 [95%CI 1.04-1.11], p<0.001; FiO_2_: OR 35.98 [95%CI 8.67-158.60], p<0.001, VIF<1.11).

**Conclusions:** PaO2-levels in patients with ARDS and VV-ECMO have a U-shaped association with mortality. Optimal outcomes are observed in the 90-123 mmHg range, which is higher compared to non-ECMO settings. Whether this is explainable by increased tissue oxygenation with concurrent avoidance of pulmonary hypoxia should be subject of future research.

**Research Letter:** Optimal oxygen target levels for critically ill patients requiring mechanical ventilation have been discussed for decades and recent guidelines recommend PaO_2_-levels between 70 and 90 mmHg.^1–3^ In contrast, very low but also high PaO_2_-levels were both associated with higher mortality, yielding a U-shaped association between PaO_2_ and mortality.^4–6^ In patients with ARDS and refractory hypoxemic respiratory failure, veno-venous Extracorporeal Membrane Oxygenation (VV-ECMO) can be implemented as a rescue therapy and PaO_2_-levels can be controlled by the blood flow of the VV-ECMO.^7^ Yet, optimal oxygenation targets in ARDS patients treated with VV-ECMO are unknown.

---

Patients with ARDS admitted to a tertiary ARDS referral center and treated with VV-ECMO were included in this study. Data collection was performed as previously reported from two electronic patient data management systems (SAP, Walldorf, Germany and COPRA, Sasbachwalden, Germany).^8^ The study was approved by the Medical Ethics Committee of Charité–Universitätsmedizin Berlin EA2/019/19. Time-weighted averages of PaO_2_ and FiO_2_ over VV-ECMO therapy time were calculated. Simple logistic regression analysis was performed for mortality and mean PaO_2_. Considering a possible non-linear relation between mortality and PaO_2_, a restricted cubic spline (RCS) for mean PaO_2_ was utilized for a simple logistic regression model.^9^ The knot values determined by the RCS function were further used for grouping and risk stratification in a regression model. A bivariate logistic regression model was calculated for mean PaO_2_ and mean FiO_2_ as well as the RCS of PaO_2_. For all multivariate models, collinearity was assessed, and variance inflation factors (VIF) were calculated. A two-tailed p-value of 0.05 was considered significant. All analyses were performed with R (R Project for Statistical Computing) Version 4.5.1.

A total of 443 patients with ARDS and treatment with VV-ECMO was included in the analysis. In this patient cohort, 88.328 arterial blood gas measurements taken during therapy with VV-ECMO were available. Mean PaO_2_ differed significantly between survivors and non-survivors (95.70 [87.46,109.20] vs. 84.27 [75.36, 96.99], p<0.001) as did mean FiO_2_ (0.54 [0.45-0.62] vs. 0.66 [0.54-0.81], p<0.001).

A simple logistic regression showed a significant positive association for mean PaO_2_ and ICU mortality (per mmHg OR 0.99 [95%CI 0.98-1.00], p=0.002). RCS analysis showed a U-shaped association of mortality and mean PaO_2_ (per mmHg PaO_2_ : PaO_2_ 69.70-90.24mmHg: OR 0.92 [95%CI 0.89-0.94], p<0.001, PaO_2_ 90.24-123.40mmHg: OR 1.09 [1.06-1.13], p<0.001, Figure 1) with both low and high PaO_2_-levels associated with an increased risk of death. Using the identified values for risk stratification revealed a significant risk reduction for patients with a mean PaO_2_ over 90.24mmHg (PaO_2_ 90.24-123.40mmHg: OR 0.25 [0.16-0.38], p<0.001; PaO_2_>123.40mmHg: OR 0.43 [0.22-0.84], p=0.0131) compared to patients with a mean PaO_2_ 69.70-90.24mmHg (OR 1 (reference group)), indicating a survival benefit for patients on higher PaO_2_ levels. Mortality did, however, not increase further with even lower PaO_2_-levels, as no significant risk difference was detected for patients with mean PaO_2_-values below 69.70 (PaO_2_ <69.70: OR 1.33 [0.66-2.79], p=0.437) compared to the reference group (PaO_2_ 69.70-90.24mmHg).

**Figure 1.**
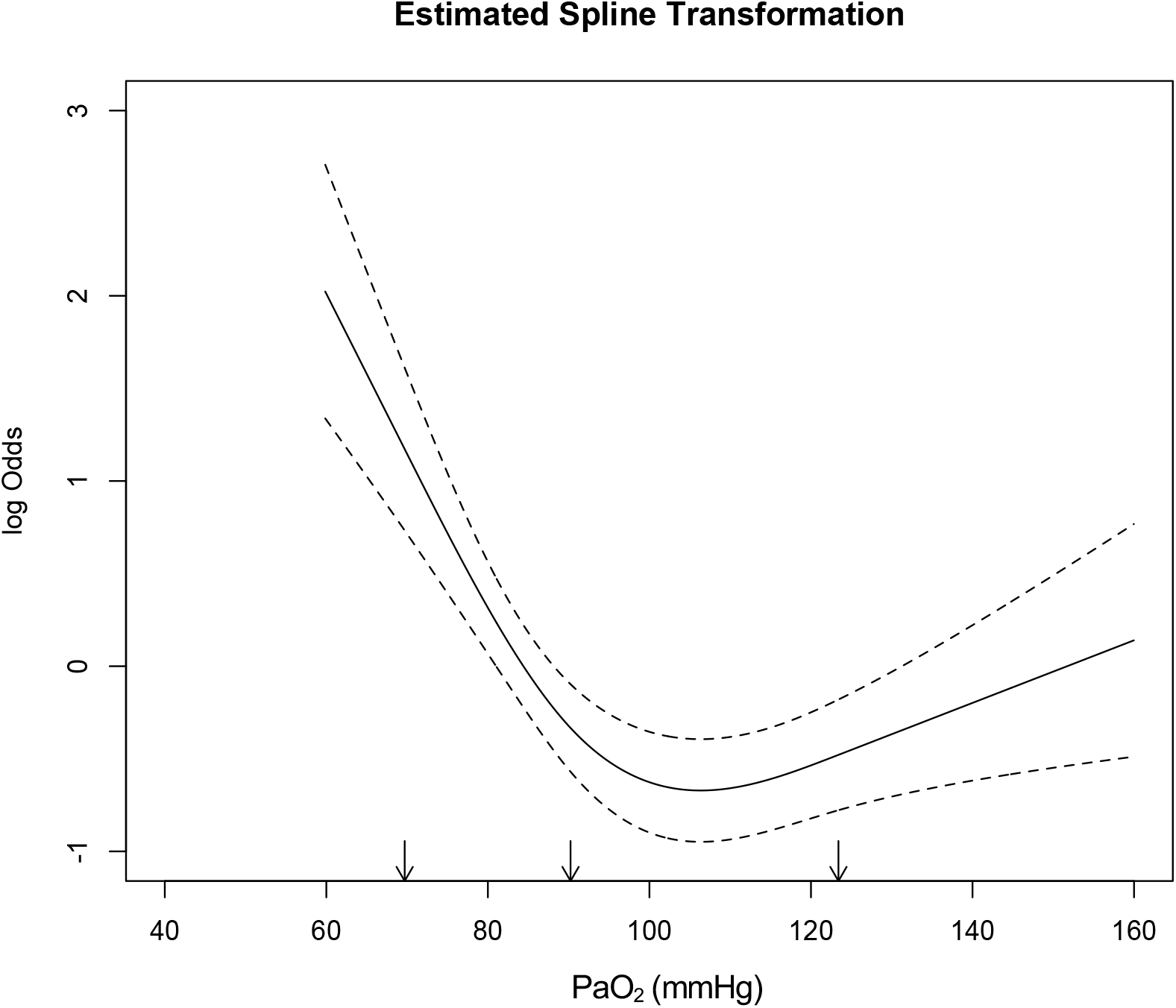
Restricted cubic spline regression model of PaO_2_ and mortality. Arrows indicate the knots at PaO_2_ of 69.70mmHg, 90.24mmHg and 123.40mmHg.

In the multivariable logistic regression analysis evaluating mortality predictors, PaO_2_ demonstrated no significant association with mortality (OR 1.00, 95% CI 0.99-1.01, p=0.413), whereas FiO_2_ emerged as an independent predictor of mortality (OR 82.59, 95% CI 21.19-348.68, p<0.001; Fig. 2A). Multicollinearity assessment confirmed minimal interdependence between these variables (VIF <1.11). However, a model including PaO_2_ as restricted cubic spline variable and FiO_2_ showed significant associations of mortality with both variables (PaO_2_ 69.70-90.24mmHg: OR 0.94 [0.91-0.97], p<0.001; PaO_2_ 90.24-123.40mmHg: OR 1.07 [1.04-1.11], p<0.001; FiO_2_ : OR 35.98 [8.67-158.60], p<0.001, VIF<1.11). A visualization of the predictions of the multivariable model is shown in Figure 2B.

**Figure 2.**
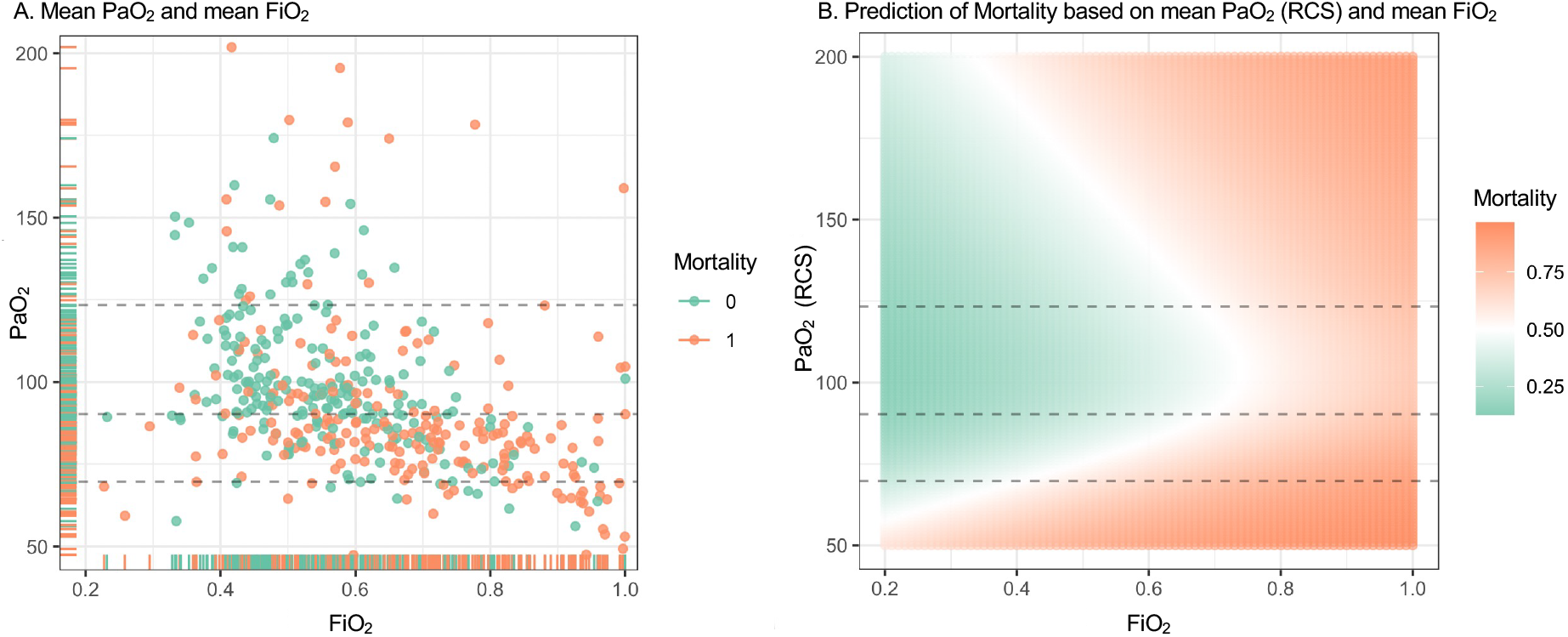
A.Mean PaO_2_ and mean FiO_2_ values colored by mortality – green – survived, orange – deceased. PaO_2_ values of knots calculated by a restricted cubic spline model (RCS) are presented as dotted lines. B. Prediction of mortality based on regression model with mean PaO_2_ (RCS) and mean FiO_2_.

In patients with ARDS requiring treatment with VV-ECMO the association between PaO_2_ and mortality follows a U-shaped curve rather than a linear trend with the lowest risk of death in the group with a PaO_2_ between 90 and 123 mmHg. This PaO_2_-range is higher compared to non-ECMO settings.

Patients with ARDS differ from the general ICU population in their reduced potential to achieve extreme hyperoxemia. Similar to the results of the present study, Boyle and colleagues demonstrated in 202 patients with ARDS that PaO_2_-levels during the first 7 days of ARDS and mortality had a U-shaped relationship.^5^ The lowest ICU-mortality risk was detected when the average time-weighted PaO_2_ was between 93.8 and 105 mmHg, a target range identical to the liberal PaO_2_ target group in the LOCO_2_ study.^10^ In patients treated with a VV-ECMO, PaO_2_-levels can be controlled by the blood flow of the VV-ECMO facilitating improvement of blood oxygenation and tissue supply without the need for exposure of the lung to excessive FiO_2_ and associated hyperoxic lung injury. Increased tissue oxygenation with concurrent avoidance of pulmonary hypoxia might be a mechanism for the reduced mortality in patients with ARDS and VV-ECMO exposed to higher PaO_2_-levels. To our knowledge, this is the first study analyzing PaO_2_-target values exclusively in patients with ARDS and during treatment with VV-ECMO.

This study is limited by the retrospective and single-center design potentially limiting generalizability to different patient populations or management approaches. The observational nature precludes definitive causal inferences regarding the relationship between PaO_2_-levels and mortality outcomes. Sicker patients might have received higher FiO_2_ due to operator bias. Despite our rigorous analytical approach, unknown and/or unmeasured confounding variables may influence the observed associations. In addition, the potential interaction between PaO_2_-levels and other physiological parameters such as arterial carbon dioxide tension, pH, hemoglobin concentration, or cardiac output, which collectively determine oxygen delivery to tissues was not analyzed. Finally, very short periods of relevant hypoxemia have a tremendous impact on mortality which is neglected analyzing time-weighted averages of the PaO_2_ - and FiO_2_-levels.

This study demonstrates that PaO_2_-levels in patients with ARDS and treatment with VV-ECMO have a U-shaped association with mortality. Optimal outcomes are observed in the 90-123 mmHg range, which is higher compared to non-ECMO settings. FiO_2_ requirements remain strongly associated with mortality independent of the achieved PaO_2_-levels, suggesting that both parameters should be considered in oxygenation management strategies. Whether increased tissue oxygenation with concurrent avoidance of pulmonary hypoxia is beneficial should be subject of future research.

## Data Availability

Data are available from the corresponding author on reasonable request.

## Declarations

### Ethics approval and consent to participate

Patient consent was waived due to the retrospective nature of the study (Ethical committee of Charité – Universitätsmedizin Berlin: No. EA2/019/19).

### Availability of data and materials

Data are available from the corresponding author on reasonable request.

## Acknowledgements

Dr Bünger is supported by the Rahel Hirsch Programme of Charité – Universitätsmedizin Berlin. Dr Bünger and Luigi La Via are participants in the ESAIC Mentorship Programme (ESAIC_MSP_2024_VB).

## Abbreviations

ICU: intensive care unit
ARDS: acute respiratory distress syndrome
ECMO: extracorporeal membrane oxygenation
paO_2_: partial pressure of oxygen
FiO_2_: fraction of inhaled oxygen
PEEP: positive end-expiratory pressure
RCS: restricted cubic spline

